# Recent UK type 2 diabetes treatment guidance represents a near whole population indication for SGLT2-inhibitor therapy

**DOI:** 10.1101/2023.08.04.23293666

**Authors:** Katherine G Young, Rhian Hopkins, Beverley M Shields, Nicholas J Thomas, Andrew P McGovern, John M Dennis

## Abstract

Recent type 2 diabetes guidance from the UK’s National Institute for Health and Care Excellence (NICE) proposes offering SGLT2-inhibitor therapy to people with established atherosclerotic cardiovascular disease (ASCVD) or heart failure, or at high-risk of cardiovascular disease defined as a 10-year cardiovascular risk of >10% using the QRISK2 algorithm.

We used a contemporary population-representative UK cohort of people with type 2 diabetes to assess the implications of this guidance. 93.1% of people currently on anti-hyperglycaemic treatment are now recommended or considered for SGLT2-inhibitor therapy, with the majority (59.6%) eligible on the basis of QRISK2 rather than established ASCVD or heart failure (33.6%). Applying these results to the approximately 2.20 million people in England currently on anti-hyperglycaemic medication suggests 1.75 million people will now be considered for additional SGLT2-inhibitor therapy.

Given older people, those of non-white ethnic groups, and those at lower cardiovascular disease risk were underrepresented in the clinical trials upon which this guidance was based, careful evaluation of the impact and safety of increased SGLT2-inhibitor prescribing across different populations is urgently required. Evidence of benefit should be weighed against the major cost implications for the NHS.

---

Recent type 2 diabetes guidance from the UK’s National Institute for Health and Care Excellence (NICE) proposes offering SGLT2-inhibitor therapy to people with established atherosclerotic cardiovascular disease (ASCVD) or heart failure, or at high-risk of cardiovascular disease defined as a 10-year cardiovascular risk of >10% using the QRISK2 algorithm.(1) The guidance applies irrespective of glycaemic control and both for people initiating first-line anti-hyperglycaemic treatment and those established on therapy. By including those without established ASCVD, this guidance goes beyond the evidence from clinical trials of the cardiovascular benefit of SGLT2-inhibitors in which a majority of participants (57-85%) had established cardiovascular disease.(2) The implications of this proposal on national prescribing are not reported.

We used a contemporary population-representative UK cohort of people with type 2 diabetes (n=568,524) actively registered with a GP practice in February 2020 (using Clinical Practice Research Datalink) to assess the implications of this guidance. 93.1% of people currently on anti-hyperglycaemic treatment are now recommended or considered for SGLT2-inhibitor therapy, with the majority (59.6%) eligible on the basis of QRISK2 rather than established ASCVD or heart failure (33.6%) (Figure 1). Prescribing of SGLT2i at the time of evaluation was limited to 14.9% of people on anti-hyperglycaemic treatment, of whom a minority had established ASCVD or heart failure (26.6%).

**Figure 1:**
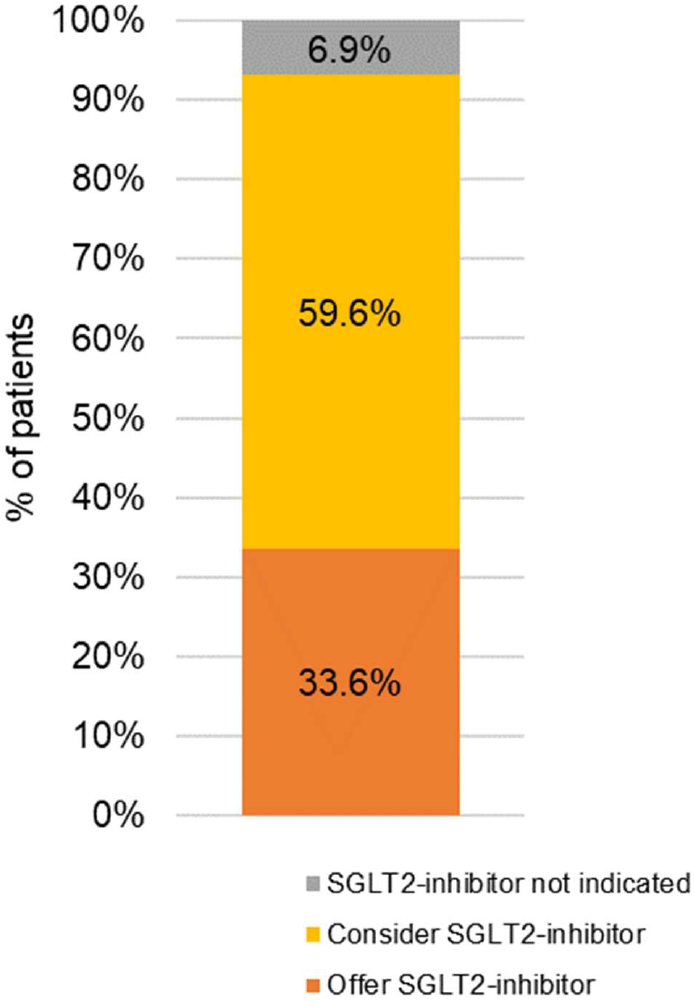
SGLT2-inhibitor therapy recommendations for people with type 2 diabetes currently treated with anti-hyperglycaemic medication in UK primary care. Estimates are derived from a UK cohort of people with type 2 diabetes (Clinical Practice Research Datalink, n=568,524 actively registered with a GP practice in February 2020). People with chronic kidney disease stage 4-5 were excluded (n=18,594) as this group represents a specific population with different criteria for SGLT2-inhibitor initiation. Remaining individuals were classified as either currently receiving anti-hyperglycaemic treatment (73.0%, n=415,267) or currently untreated (27.0%, n=153,257). NICE criteria was then used to classify individuals by cardiovascular disease status as 1) having established atherosclerotic cardiovascular disease or heart failure [Offer SGLT2-inhibitor]; 2) at high-risk of cardiovascular disease as defined by a 10-year risk of cardiovascular disease >10% applying the QRISK2 algorithm [Consider SGLT2-inhibitor]; 3) not at high-risk of cardiovascular disease (QRISK2 <10%) [SGLT2-inhibitor not indicated]. Amongst the 27.0% currently untreated individuals, the majority of whom will eventually require treatment, 93.3% are recommended or considered for SGLT2-inhibitor therapy, with the majority (57.6%) eligible on the basis of QRISK2 rather than established ASCVD (35.7%) [data not shown].

Applying these results to the approximately 2.20 million people in England currently on anti-hyperglycaemic medication suggests 1.75 million people will now be considered for additional SGLT2-inhibitor therapy. Full implementation could increase the cost of SGLT2-inhibitor prescribing from £147 million(3) to over £900 million per year. For context, total annual spending on cholesterol-lowering drugs in the whole population with and without diabetes is around £215 million.(4)

The updated NICE guidance represents a near population-level intervention of SGLT2-inhibitor initiation for people with type 2 diabetes. Recommendations are based on extrapolation of trial evidence to people at lower cardiovascular risk. Given older people and non-white ethnic groups are under-represented in trials, careful evaluation of the impact and safety of increased SGLT2-inhibitor prescribing across different populations is urgently required. Evidence of benefit should be weighed against the major cost implications for the NHS. If guidelines are adhered to, increases in SGLT2-inhibitor prescribing could near double the £1 billion annual spend on diabetes drug prescribing.(5)

## Data Availability

CPRD data are available by application, subject to protocol approval through CPRD's Research Data Governance (RDG) Process.

## Acknowledgements

This article is based in part on data from the Clinical Practice Research Datalink (CPRD) obtained under licence from the UK Medicines and Healthcare products Regulatory Agency (ISAC 20_000101). CPRD data is provided by patients and collected by the NHS as part of their care and support.

## Contributor and guarantor information

The concept and design were developed by APM, JMD, KGY and NJT. KGY and RH undertook the analysis, with support from JMD, APM, NJT and BMS. All authors provided support for the analysis and interpretation of results, critically revised the article, and approved the final article. JMD attests that all listed authors meet authorship criteria and that no others meeting the criteria have been omitted.

